# Transmission prevention behaviors in US households with SARS-CoV-2 cases in 2020

**DOI:** 10.1101/2022.11.25.22282730

**Authors:** Rebecca Rubinstein, Wenwen Mei, Caitlin A. Cassidy, Gabrielle Streeter, Christopher Basham, Carla Cerami, Feng-Chang Lin, Jessica T. Lin, Katie R. Mollan

**Affiliations:** Department of Epidemiology, University of North Carolina at Chapel Hill Gillings School of Global Public Health, Chapel Hill, NC, USA; Department of Biostatistics, University of North Carolina at Chapel Hill Gillings School of Global Public Health, Chapel Hill, NC, USA; University of North Carolina at Chapel Hill, Chapel Hill, NC, USA; Institute of Global Health and Infectious Diseases, University of North Carolina at Chapel Hill School of Medicine, Chapel Hill, NC, USA; Medical Research Council Unit, The Gambia, at the London School of Hygiene & Tropical Medicine, The Gambia; Center for AIDS Research, University of North Carolina at Chapel Hill School of Medicine, Chapel Hill, NC, USA

## Abstract

**Background:** SARS-CoV-2 transmission frequently occurs within households, yet few studies describe which household contacts and household units are most likely to engage in transmission-interrupting behaviors.

**Methods:** We analyzed a COVID-19 prospective household transmission cohort in North Carolina (April-Oct 2020) to quantify changes in physical distancing behaviors among household contacts over 14 days. We evaluated which household contacts were most likely to ever mask at home and to ever share a bedroom with the index case between Days 7-14.

**Results:** In the presence of a household COVID-19 infection, 24% of household contacts reported ever masking at home during the week before study entry. Masking in the home between Days 7-14 was reported by 26% of household contacts, and was more likely for participants who observed their household index case wearing a mask. Participants of color and participants in high-density households were more likely to mask at home. After adjusting for race/ethnicity, living density was not as clearly associated with masking. Symptomatic household contacts were more likely to share a bedroom with the index case. Working individuals and those with comorbidities avoided sharing a bedroom with the index case.

**Conclusion:** In-home masking during household exposure to COVID-19 was infrequent in 2020. In light of ongoing transmission of SARS-CoV-2, these findings underscore a need for health campaigns to increase the feasibility and social desirability of in-home masking among exposed household members. Joint messaging on social responsibility and prevention of breakthrough infections, reinfections, and long COVID-19 may help motivate transmission-interruption behaviors.

## INTRODUCTION

Households are a high-risk setting for transmission of Severe Acute Respiratory Syndrome Coronavirus-2 (SARS-CoV-2), especially when SARS-CoV-2-positive individuals are unable to self-isolate. Infected individuals may face challenges distancing from family members and wearing masks at home, and they are unlikely to take precautions just prior to symptom onset, when viral shedding and infectiousness peak.[1–4] In 2020, before widespread vaccination, high household secondary attack rates were identified in the US, including a rate of 52% among households in Wisconsin and Tennessee and 60% in North Carolina.[4,5] A majority of secondary cases were identified within a week of the index case presenting symptoms.[4,5] Although vaccination greatly reduces the likelihood of severe disease, outbreaks of the more-transmissible Delta and Omicron variants and sub-variants have occurred among vaccinated index cases and close contacts in households across the US.[6–8]

Modifiable risk factors to help interrupt household transmission include masking at home, and avoiding sharing a bedroom with infected individuals.[4,8,9] Previous studies support immediate isolation within one’s household upon testing positive.[8] However, few published studies have characterized which household contacts and household units are most likely to engage in behaviors that interrupt transmission, and the structural barriers that can prevent them from doing so, including high household living density.[10,11]

The aims of the current study are 1) to describe changes in household contacts’ COVID-19 mitigating behaviors (e.g., mask-wearing, sharing a bedroom with primary infected case) between cohort entry and Day 14 of cohort participation and 2) to identify structural and individual-level factors associated with these behaviors at Day 14. We analyzed behavioral data from the COVID-19 Household Transmission Study (CO-HOST), a racially and ethnically diverse cohort of household transmission in central North Carolina conducted from April to October 2020, encompassing rural, suburban and urban households.[4] In 2020, both the original Wuhan strain of SARS-CoV-2 and the D614G “G” variant circulated across the US.[12] At that time, public health guidance recommended 14 days of self-quarantine following possible COVID exposure. Our findings can help guide prevention efforts for household transmission of SARS-CoV-2 in North Carolina and comparable regions. Given the frequency of novel and highly-transmissible SARS-CoV-2 variants and challenges to herd immunity in the US,[13,14] including vaccine hesitancy,[15] a better understanding of behaviors that contribute to preventing transmission in infected households can alleviate future waves of SARS-CoV-2 in the US.

## METHODS

### Study sample and design

The CO-HOST study recruited patients infected with SARS-CoV-2 who tested at a UNC Respiratory Diagnostic Center in Chapel Hill, Cary or Raleigh, NC (index cases). Adults testing positive for SARS-CoV-2 were recruited with their household members (household contacts) over 2 years of age, who planned to spend at least 4 weeks in the same house as the index case. The primary aim of CO-HOST was to determine the household secondary attack rate of SARS-CoV-2 infection in central North Carolina. Detailed inclusion criteria, follow-up testing, classification of index cases and household contacts, and study aims have been previously described.[4] Ethical approval for the parent study was received from the Institutional Review Board at the University of North Carolina at Chapel Hill (Protocol Number 20-0982), participants gave informed consent before participating, and the parent study conformed to the principles outlined in the Declaration of Helsinki.

At cohort entry (Day 0), along with PCR nasopharyngeal and saliva testing, we asked all index cases and household contacts whether they ever masked at home in the previous 7 days. Participants were also asked about COVID-19 symptoms, comorbidities, sociodemographic characteristics, and their activities in the prior week. They completed electronic symptom diaries until 2 consecutive days without symptoms or until day 21 if they never developed symptoms. If participants missed ≥2 days of questionnaires, symptoms were ascertained by study staff over the phone.[4] At Day 14, household contacts again received testing and answered the same questions asked at baseline.

### Outcomes

Household contacts were asked whether they engaged in the following activities with the index case at cohort entry and Day 14: sharing bedroom, sharing bathroom, sharing kitchen, watching television, eating together, sharing car rides, and sharing electronic devices. The primary behavioral outcomes for inferential analyses were 1) did the household contact ever wear a mask at home between days 7 and 14 (yes/no) and 2) did the household contact ever share a bedroom with the index case between days 7 and 14 (yes/no).

### Exposures

We assessed the association of the following individual-level factors to the outcomes: age, sex, race/ethnicity, and aged 50 or older/reporting ≥1 comorbidity. We also assessed the following factors 7-14 days after cohort entry: COVID-19 symptom duration, primary caregiving to the index case, and working outside the home. For each household contact, we assessed household-level exposures including high living density (>3 individuals in <6 rooms, including bedrooms, kitchen, and common rooms, but not bathrooms or garage) and whether the household contact observed the index case wearing a mask 7-14 days after cohort entry.

### Statistical analysis

Fourteen-day changes in the proportion of household contacts engaged in shared behaviors with the index case were estimated among participants with non-missing responses. To account for clustering within households, we used the Yang modification of Obuchowski’s test for changes in paired binary data[16], executed in the clust.bin.pair package (v01.1.2) of R version 4.0.5.[17]

We estimated associations between exposure variables and household contacts 1) ever masking at home and 2) sharing a bedroom with the index case at Day 14 using log-binomial models fit with generalized estimating equations (GEE) to account for clustering of contacts within households (using Windows SAS 9.4). For each outcome, intra-cluster correlation (ICC) was estimated from an intercept-only model fit with GEE using an exchangeable working correlation. In sensitivity analyses, missing data were handled using multiple imputation (MI) for clustered multi-level data, using the jomo package in R version 4.0.2.[18,19] A type I error rate of alpha 0.05 was applied throughout, with no adjustment for multiplicity.

## RESULTS

Between April and October 2020, 100 households with 204 eligible household contacts were enrolled into CO-HOST.[4] Two households and 4 household contacts were excluded due to incomplete study follow-up (Figure S1). A majority of household contacts did not know their own infection status while answering surveys at cohort entry and Day 14, although they were aware that the index case was infected at study entry. Despite not necessarily knowing their own infection status, over half (54%) of household contacts at cohort entry reported symptoms consistent with COVID-19 infection in the previous 7 days (Table 1).

**Table 1.** Characteristics of household contacts at cohort entry.

| **Variable** | **Overall** | **BIPOC<sup>a</sup>** | **White, non-Hispanic** |
| --- | --- | --- | --- |
| **Household-level characteristics** | **N=100 households** | **N=54 households** | **N=46 households** |
| Number of household members in each household |  |  |  |
| 2 people | 27 (27.0) | 14 (25.9) | 13 (28.3) |
| 3 people | 23 (23.0) | 11 (20.4) | 12 (26.1) |
| 4 people | 22 (22.0) | 9 (16.7) | 13 (28.3) |
| 5 or more people | 28 (28.0) | 20 (37.0) | 8 (17.4) |
| Number of rooms in house<sup>b</sup> |  |  |  |
| 2 or fewer rooms | 10 (10.0) | 7 (13.0) | 3 (6.5) |
| 3-5 rooms | 43 (43.0) | 31 (57.4) | 12 (26.1) |
| 6 or more rooms | 47 (47.0) | 16 (29.6) | 31 (67.4) |
| Number of square feet in house |  |  |  |
| <500 sq feet (<46.5 sq m) | 3 (3.0) | 2 (3.7) | 1 (2.2) |
| 500-1000 sq feet (46.5-93 sq m) | 17 (17.0) | 12 (22.2) | 5 (10.9) |
| 1000-2000 sq feet (93-186 sq m) | 33 (33.0) | 19 (35.2) | 14 (30.4) |
| >2000 sq feet (>186 sq m) | 42 (42.0) | 16 (29.6) | 26 (56.5) |
| Unknown | 5 (5.0) | 5 (9.3) | 0 (0.0) |
| Household with high living density<sup>c</sup> |  |  |  |
| Yes | 23 (23.0) | 20 (37.0) | 3 (6.5) |
| No | 77 (77.0) | 34 (63.0) | 43 (93.5) |
| % of household members (including index cases) with COVID-like symptoms by Day 7<sup>de</sup> |  |  |  |
| <50% | 14 (22.6) | 7 (21.2) | 7 (24.1) |
| 50 to <100% | 15 (24.2) | 8 (24.2) | 7 (24.1) |
| 100% (all members) | 33 (53.2) | 18 (54.6) | 15 (51.7) |
| Missing | 4 | 2 |  |
| **Individual-level characteristics** | **N=204 participants** | **N=97 participants** | **N=107 participants** |
| Age |  |  |  |
| 0-12y | 46 (22.6) | 23 (23.7) | 23 (21.5) |
| 13-17y | 24 (11.8) | 12 (12.4) | 12 (11.2) |
| 18-24y | 25 (12.3) | 11 (11.3) | 14 (13.1) |
| 25-49y | 67 (32.8) | 35 (36.1) | 32 (29.9) |
| 50-64y | 30 (14.7) | 10 (10.3) | 20 (18.7) |
| >65y | 12 (5.9) | 6 (6.2) | 6 (5.6) |
| Current Sex |  |  |  |
| Male | 98 (48.9) | 46 (47.4) | 52 (48.6) |
| Female | 106 (52.0) | 51 (52.6) | 55 (51.4) |
| Race/Ethnicity |  |  |  |
| *White, non-Hispanic* | 107 (52.5) |  |  |
| *Hispanic/Latinx* | 70 (34.3) |  |  |
| *Black, non-Hispanic* | 18 (8.8) |  |  |
| *Other Race/Unknown Race<sup>f</sup>, non-Hispanic* | 9 (4.4) |  |  |
| **Education** |  |  |  |
| *Children under 18* | 70 (35.0) | 35 (37.6) | 35 (32.7) |
| *Adult, high-school or less* | 63 (31.5) | 41 (44.1) | 22 (20.6) |
| *College degree* | 38 (19.0) | 11 (11.8) | 27 (25.2) |
| *Graduate degree* | 29 (14.5) | 6 (6.5) | 23 (21.5) |
| *Missing* | 4 |  |  |
| **Any comorbidities<sup>g</sup>** |  |  |  |
| *Yes* | 71 (35.5) | 38 (40.0) | 33 (31.4) |
| *No* | 129 (64.5) | 57 (60.0) | 72 (68.6) |
| *Missing* | 4 | 2 | 2 |
| **BMI $\geq 30^h$** |  |  |  |
| *Yes* | 46 (31.9) | 27 (46.6) | 19 (22.1) |
| *No* | 98 (68.1) | 31 (53.5) | 67 (77.9) |
| *Missing* | 18 | 17 | 1 |
| **COVID-19 like symptoms in past 7 days<sup>d</sup>** |  |  |  |
| *Yes* | 109 (54.0) | 49 (51.6) | 60 (56.1) |
| *No* | 93 (46.0) | 46 (48.4) | 47 (43.9) |
| *Missing* | 2 | 2 | 0 |
| **Relationship to primary infected case** |  |  |  |
| *Partner* | 58 (28.7) | 21 (22.1) | 37 (34.6) |
| *Child* | 68(33.7) | 32 (33.7) | 36 (33.6) |
| *Sibling, including in-laws* | 19 (9.4) | 13 (13.7) | 6 (5.6) |
| *Parent, including in-laws* | 35 (17.3) | 19 (20.0) | 16 (15.0) |
| *Roommate/friend* | 15 (7.4) | 7 (7.4) | 8 (7.5) |
| *Other relative/other* | 7 (3.5) | 3 (3.2) | 4 (3.7) |
| *Missing* | 2 | 2 |  |
| **Caregiver to primary infected case<sup>h</sup>** |  |  |  |
| *Yes* | 58 (38.7) | 21 (31.8) | 37 (44.1) |
| *No* | 92 (61.3) | 45 (68.2) | 47 (56.0) |
| *Missing* | 12 | 9 | 3 |
| **Index case ever wore mask in the home past 7 days** |  |  |  |
| *Yes* | 153 (80.1) | 79 (90.8) | 74 (71.2) |
| *No* | 38 (19.9) | 8 (9.2) | 30 (28.9) |
| *Missing* | 13 | 10 | 3 |
| **Live with someone under 18<sup>j</sup>** |  |  |  |
| *Yes* | 72 (50.4) | 31 (49.2) | 41 (51.3) |
| *No* | 71 (49.7) | 32 (50.8) | 39 (48.8) |
| **Association to healthcare facility<sup>jk</sup>** |  |  |  |
| *Works in a healthcare facility* | 7 (5.0) | 2 (3.2) | 5 (6.3) |
| *Household includes someone who works in a healthcare facility* | 18 (12.8) | 6 (9.7) | 12 (15.2) |
| *Neither* | 116 (82.3) | 54 (87.1) | 62 (78.5) |
| *Missing* | 2 | 1 | 1 |

a Includes non-Hispanic Black, Hispanic/Latinx of any race, Asian American and Pacific Islander, Native American and Alaska Native, Other race, and Mixed race. Households were considered to be BIPOC if at least one CO-HOST participant (index case or household contact) self-identified as BIPOC.
b Including bedrooms, kitchen, and common rooms, but not bathrooms or garage
c >3 persons occupying <6 rooms, including bedrooms, kitchen, and common rooms, but not bathrooms or garage
d Symptoms assessed in daily symptom surveys included fever, chills, muscle aches, runny nose, sore throat, loss of taste or smell, cough, shortness of breath, chest pain, wheezing, nausea, diarrhea, headache, abdominal pain.
e N=66 households in which every member of the household was enrolled.
f Self-identified races include American Indian or Alaska Native, Asian, Native Hawaiian or Other Pacific Islander, Other Race, Unknown Race, or Refusal
g Comorbidities include HIV, chronic lung disease (e.g. emphysema, COPD), asthma, daily smoking, heart disease (e.g. previous heart attack, heart failure, stents), morbid obesity (>100 pounds over ideal weight), diabetes, high blood pressure, chronic kidney disease, chronic liver disease, weak immune system due to disease or medication, and recent (within past 2 weeks) or current pregnancy.
h Includes participants ages 12 and over
i Other relationships include sibling, parent, other relative, roommate and friend/non-roommate

CO-HOST household contacts were racially and ethnically diverse. Almost half (48%) of the participants self-identified as Black, Indigenous, or People of Color (BIPOC), including a high proportion of Hispanic/Latinx participants (34%). Twenty-three percent of participants resided in ‘**high**-density’ households, with more than 3 people occupying fewer than 6 living spaces (Table 1). Most participants (86%) lived with at least one other person at high risk of experiencing complications from COVID-19 infection, including individuals 50 and older and those with obesity or comorbidities. Together, these characteristics illustrate a cohort of exposed household members vulnerable to the downstream effects of COVID-19 infection. Baseline characteristics are shown separately for BIPOC and White non-Hispanic participants (Table 1).

We first assessed changes in household contact behavior from cohort entry to Day 14 (Figure 1; Table S1). Several space-sharing behaviors declined from cohort entry to Day 14, including the proportion of household contacts who shared a bedroom (36% vs. 27%, p≤0.02 or kitchen (91% vs. 76%, p≤0.003) with the index case. The proportions who ate with the index case (68% vs. 55%, p≤0.02) or rode in a car with the index case (62% vs. 41%, p≤0.001) also declined. Still, most contacts shared a kitchen (76%) or bathroom (56%) with the index case and ate or watched TV with (55% each) the index case between Days 7-14. Despite the prevalence of sharing indoor spaces, only 24% and 26% of household contacts reported that they ever masked at home at cohort entry and Day 14, respectively (Figure 1; Table S1).

**Figure 1.**
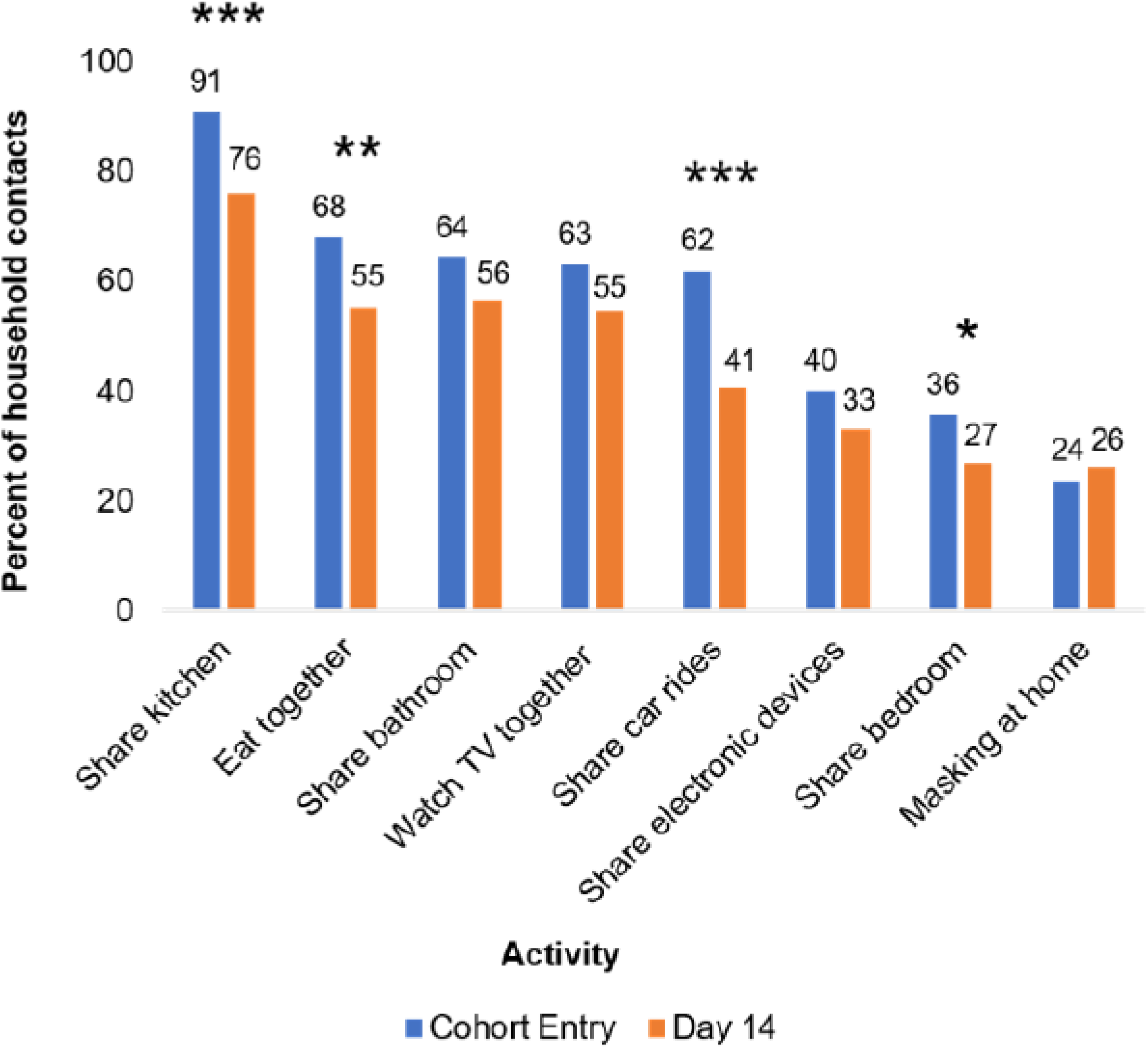
Changes in Household Contact Behaviors from Cohort Entry to Day 14. Entry encompasses the 7 days prior to cohort entry plus the day of enrollment. Day 14 encompasses Days 7-14 of participation in the cohort. Participants with non-missing data at both cohort entry and Day 14 were included in analysis. Prevalence of behaviors at each time point listed above bars. * denotes p≤0.05, ** denotes p≤0.01, and *** denotes p≤0.001. P-values were calculated using Yang’s test for changes from Day 0 to Day 14 on complete cases (Table S1). 24 and 74 participants were missing ‘masking at home’ responses at Day 0 and Day 14 respectively, and 41 participants were missing responses for all other variables.

We also assessed individual and household-level factors associated with 1) ever masking at home and 2) ever sharing a bedroom with the index case between Days 7-14. Intra-household correlation was high for the masking variable and the bedroom variable (ICC of 0.81 and 0.60, respectively). Seventy-four of 204 household contacts were missing masking data (36%) and 41 of 204 household contacts were missing bedroom data (20%) among Days 7-14.

Household contacts who self-identified as BIPOC were more likely to report masking between Days 7-14 than White, non-Hispanic contacts (Prevalence Ratio [PR]=2.0, 95% CI 1.1, 3.6). Multiple imputation (MI) did not change the strength of this association (PR=2.0, 95% CI 1.1, 3.8). Household contacts who observed the index case masking between days 7-14 were also more likely to mask at home (PR=2.0, 95% CI 1.2, 3.4). This association largely persisted in the MI analysis (PR=2.0, 95% CI 0.9, 4.2) (Figure 2; Figure S2). Contacts with longer symptom duration were also more likely to mask at home in complete case analyses (PR=1.9, 95% CI 1.0, 3.6), although this relationship did not persist in MI (PR=1.1, 95% CI 0.6, 2.0).

**Figure 2.**
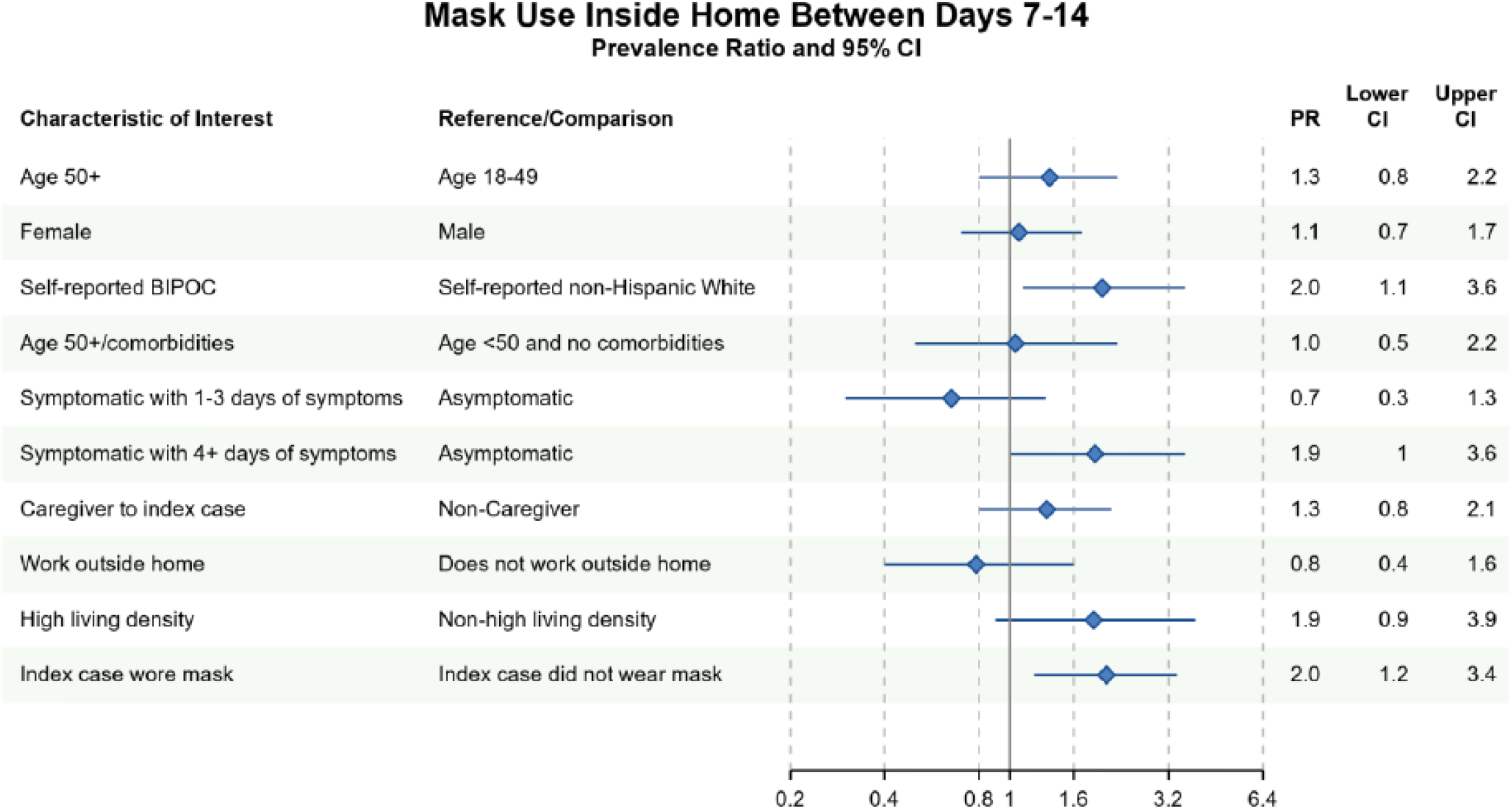
Bivariate complete case analyses of factors associated with wearing a mask at home at any time between Days7-14 of cohort participation. Dots (PR) and solid lines (95% CI) display the complete case analyses. PR and 95% CI are displayed on the natural log scale. Vertical solid line denotes the null value of the PR. X-axis labels correspond to the PR values. Sample sizes and prevalence estimates are shown in Table S4. BIPOC=Black, Indigenous, People of Color; CI=confidence interval; PR=prevalence ratio.

Different factors predicted whether household contacts shared a bedroom with the index case between Days 7-14. In both complete case and imputed analyses, household contacts were more likely to have shared a bedroom with the index case if they 1) reported 4 or more days of symptoms between days 7-14 or 2) identified as the primary caregiver to the index case between days 7-14 (Figure 3, Figure S3). Conversely, household contacts at increased risk of severe COVID-19 infection avoided sharing a bedroom with the index case in complete case (PR=0.6, 95% CI 0.4, 1.1) and imputed analyses (PR=0.7, 95% CI 0.4, 1.1). There was no evidence of an association between household contact race/ethnicity and bedroom-sharing in complete case, nor imputed sensitivity analyses (PR=1.0, 95% CI 0.6, 1.7). Similarly, index masking behavior was not associated with household contacts sharing a bedroom with the index case in neither complete case (PR=1.1, 95% CI 0.6, 2.1) nor imputed analyses (PR=1.2, 95% CI 0.7, 2.0) (Figure 3; Figure S3).

**Figure 3.**
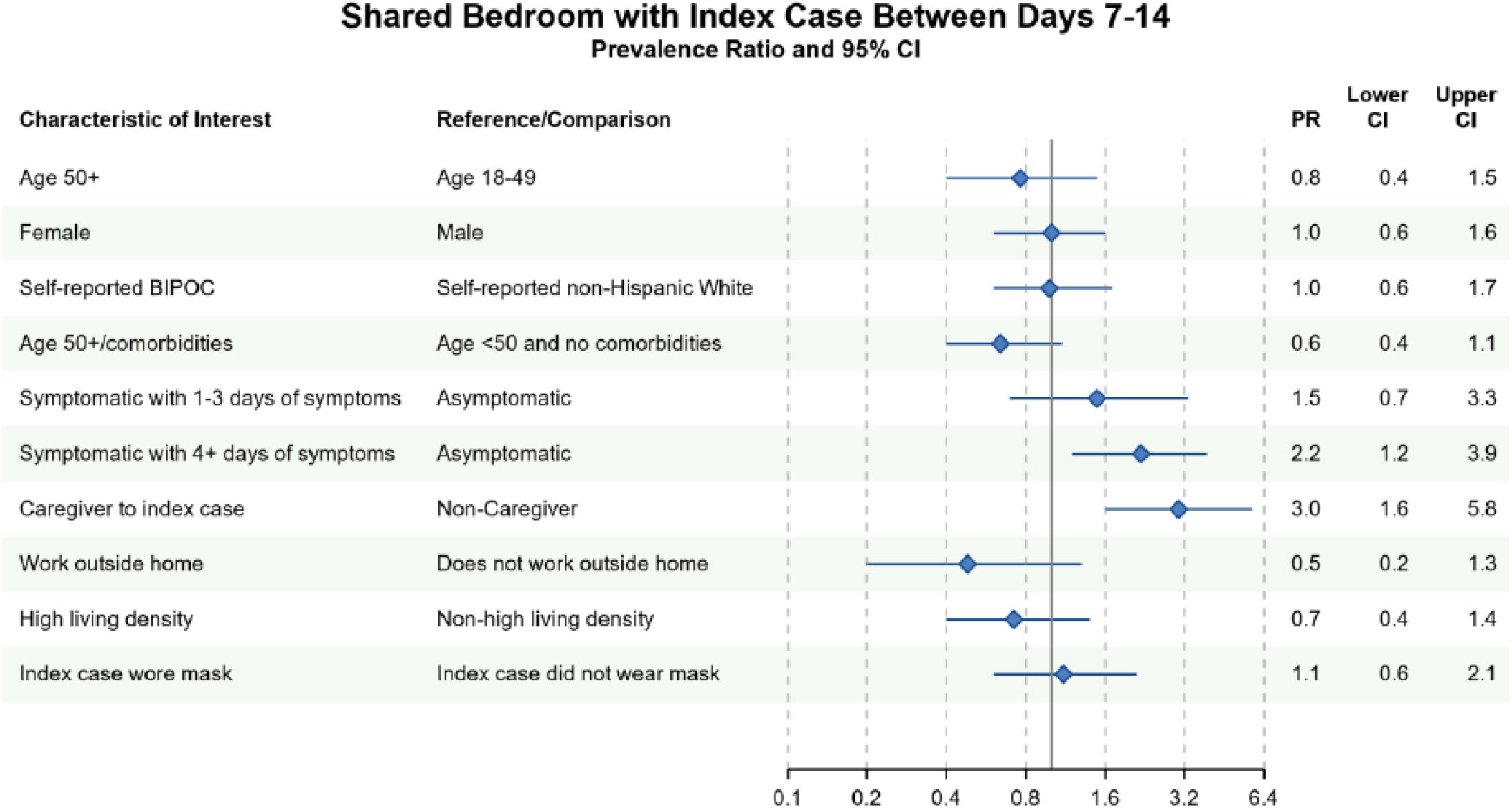
Bivariate complete case analysis of factors associated with sharing a bedroom with the index case at any time between Days 7-14 of cohort participation. Dots (PR) and solid lines (95% CI) display the complete case analyses. PR and 95% CI are displayed on the natural log scale. Vertical solid line denotes the null value of the PR. X-axis labels correspond to the PR values. Sample sizes and prevalence estimates are shown in Table S4. BIPOC=Black, Indigenous, People of Color; CI=confidence interval; PR=prevalence ratio. Table denotes the PR, lower 95% CI and upper 95% CI.

Lastly, given the associations between race/ethnicity and masking, and living density and masking, we sought to determine whether living density differed among BIPOC and White, non-Hispanic participants who masked versus those who did not. Among study participants, BIPOC were more likely than White non-Hispanics to live in a high-density-household (Table 2S, Table 3S). Among BIPOC household contacts, the likelihood of in-home masking was similar for those in a high living density household vs. a lower density household in both complete-case (PR=1.3, 95% CI 0.5, 3.1) and MI (PR= 1.2, 95% CI 0.6 2.5). The association between living in a high-density household and masking was attenuated towards the null when adjusted for BIPOC race/ethnicity in both complete case (PR=1.3, 95% CI 0.6, 3.0) and MI analysis (PR=1.2, 95% CI 0.6, 2.5).

## DISCUSSION

We prospectively examined associations between household and individual-level factors and transmission-modifying behaviors in households with active COVID-19 infections in a racially and ethnically diverse sample of North Carolina residents. Throughout the 14 days of observation, most household contacts reported not masking inside the home at any time. Nonetheless, we find that household contacts of color and contacts who observed the index case masking were much more likely to mask.

Throughout the study period, over 50% of household contacts continued to share kitchen space, share a bathroom, eat meals and watch TV with the index case. Our findings suggest that changing behaviors constrained by space and resources such as sharing bathrooms and kitchens may be difficult for households. Masking, alternatively, is an inexpensive intervention accessible to most people. Targeted demographic groups, such as White, non-Hispanic households, could be encouraged to mask more frequently, and encouraging infected or symptomatic individuals to mask at home may help convince other household members to also mask.

Unlike other studies that measured household transmission of SARS-CoV-2,[8] or behavioral interventions at the community level, our study prospectively measured the behaviors of household contacts after an initial household infection was identified. In early 2022, Baker and colleagues reported a retrospective analysis of behaviors of household members exposed to SARS-CoV-2 in Chicago, Milwaukee, Connecticut, and Utah in the winter of 2021-2022. However, their analysis did not identify demographic characteristics of household contacts who engaged in behaviors such as masking, instead focusing on behaviors associated with transmission,[8] as did our primary analysis of CO-HOST participants.[4]

Other studies evaluated attitudes and beliefs towards masking and isolating from family members if exposed to SARS-CoV-2, although they did not prospectively measure household contacts’ behavior. In the United Kingdom, adults were asked whether they would self-isolate away from home if infected or exposed if they were provided appropriate accommodations at no cost.[20] Among participants who noted that they would not be able to isolate from family members at home if infected, 56% noted that would definitely or probably accept accommodations if offered to them. Many of these individuals cited household size and the number of household residents as barriers to isolating within the home. In interviews, low-income participants and participants from racial and ethnic minority communities highlighted the elevated risk of exposure they faced at work as a driving force to accept free accommodations outside the home.

In our study, similar concerns may also explain why BIPOC household contacts and contacts living in high-density households were more likely to have masked at home, although we did not ask household contacts why they masked. While there was no clear association between living density and masking after adjusting for BIPOC race/ethnicity, BIPOC participants were overall more likely to live in high-density households. It is plausible that participants of color within our study understandably had a greater concern of contracting and surviving infection, given highly publicized racial disparities in COVID-19 infection and fatalities as early as Spring, 2020.[21,22] These concerns could have motivated BIPOC participants to mask at home, given structural barriers to isolation such as high living density, and the lack of government-sponsored accommodations for exposed or infected individuals to isolate in North Carolina and much of the US.

In our study, household contacts who observed their index case masking at home were themselves more likely to mask. Household members may share similar beliefs around the efficacy of masking, the science of SARS-CoV-2 transmission, and the severity of the virus.[8] In a ‘Prisoners’ Dilemma’ simulation of mask wearing among US adults, participants who chose not to wear masks were more likely to cooperate with non-mask wearers than mask-wearers, suggesting that in-group dynamics and social identity play a role in the decision to mask.[23] Together, findings from our study and the Prisoners’ Dilemma simulation suggest that campaigns encouraging infected and symptomatic individuals to mask at home may encourage their household members to mask as well. In-home masking may be particularly feasible for asymptomatic positive individuals, whereas some individuals with respiratory symptoms or young children may find it difficult to mask consistently. Moreover, masking is not recommended during sleep,[24] underscoring the importance of having the ability to sleep in a separate bedroom from infected individuals.

Our analyses of bedroom-sharing identified that household contacts who worked outside the home in the previous week or who had risk factors for severe COVID-19 were less likely to share a bedroom with the index case, and that individuals with 4 or more days of symptoms were more likely to have shared a bedroom with the index case. In our cohort, secondary infections were more likely among household contacts who shared a bedroom with the index case.[4] Our findings suggest that household contacts who faced steeper consequences of infection (e.g. missed days of work, higher risk of severe COVID-19) opted not to share a bedroom with the index case where possible.

Strengths of our study include the longitudinal design, a racially diverse sample, the use of multiple imputation to account for missing data, and the unique scope of our question on structural household factors associated with behaviors that affect household transmission. Our study nonetheless is limited by sample size. The masking variable was phrased as ‘ever masked at home’ versus not, which does not measure masking frequency. Additionally, while the study was prospective, behavioral outcomes were ascertained only at two timepoints. Lastly, recall bias and social desirability bias could weaken the validity of our results.

We investigated predictors of physical distancing behaviors among household contacts exposed to SARS-CoV-2 in a period of high susceptibility to COVID-19 infection. Vaccines were not available and most people were un-exposed.[25] Today, widespread vaccination and therapeutics (e.g., nirmatrelvir and ritonavir) have reduced the risk of severe disease.[26,27] However the risk of household transmission and long COVID-19 complications remains considerable,[28–30] given increased transmissibility and immune escape among new variants leading to an increase in breakthrough infections and reinfections.[31] In the ongoing phase of the COVID-19 pandemic, our findings support additional congressional funding to continue the Biden administration’s SARS-CoV-2 at-home rapid antigen test distribution program to any American household. We also encourage the administration to distribute N95 masks at the federal level, given the prohibitive cost for large households. Virtually no published studies have assessed the attitudes and motivations for masking and isolating among infected and exposed household members in the US, a large and diverse country where many communities likely have their own beliefs and barriers around masking and isolation at home. Nonetheless, we have sufficient information to justify public health campaigns increasing the feasibility and social desirability of masking and isolating among exposed household members where possible, and the need for government and private-sector support of outside accommodations where isolation and masking are impossible.

## Data Availability

De-identified participant-level data are available upon reasonable request from Rebecca Rubinstein by emailing. Please cite this manuscript upon use in further publications.

## ACKNOWLEDGEMENTS

We thank the CO-HOST study participants for their generous time and participation in the study. We are also grateful to Brian Pence, Joseé Dussault, and the UNC Center for AIDS Research Biostatistics Core for valuable input and support to improve the manuscript.

## SUPPLEMENTAL MATERIAL

**Figure S1.**
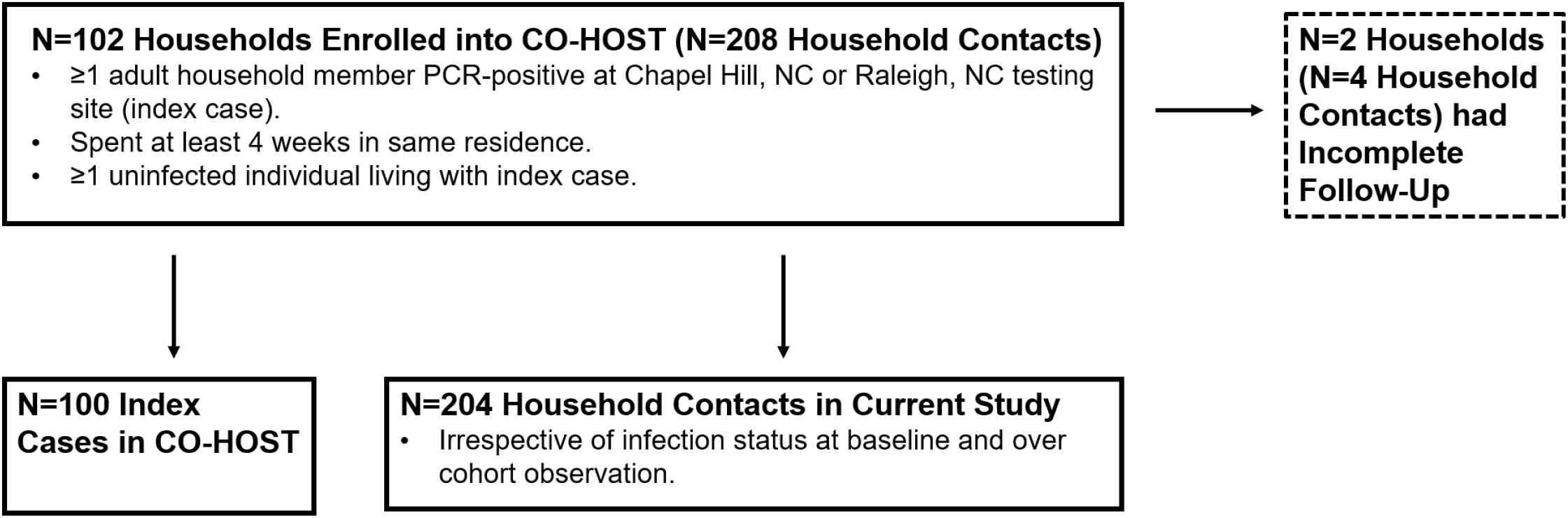
Inclusion diagram. CO-HOST refers to the parent study from which this secondary analysis is derived. 2 households consisting of 2 index cases and 4 household contacts were unable to be included in this study due to insufficient follow up. This study utilized the 204 remaining household contacts.

**Figure S2.**
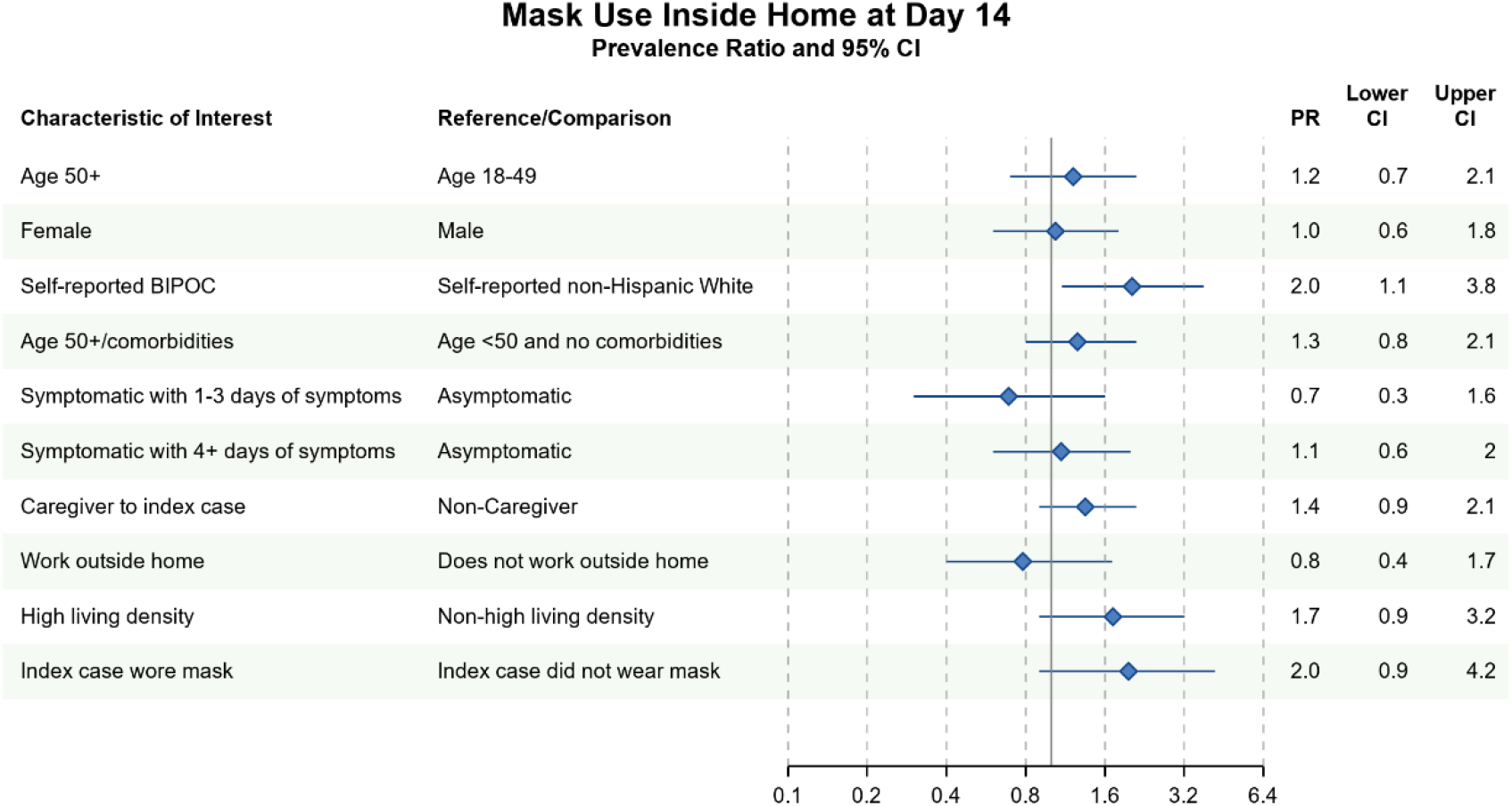
Bivariate sensitivity analyses of factors associated with using a mask at home at any time at Day 14 of cohort participation. Solid dots (PR) and solid lines (95% CI) display imputed estimates <u>using chained multiple imputation for clustered data</u>. PR and 95% CI graphed on the natural log scale. Vertical solid line denotes the null value of the PR (PR=1.0) on the exponentiated scale. X-axis labels correspond to the exponentiated scale. BIPOC=Black, Indigenous, People of Color. Table denotes the PR, lower 95% CI and upper 95% CI.

**Figure S3.**
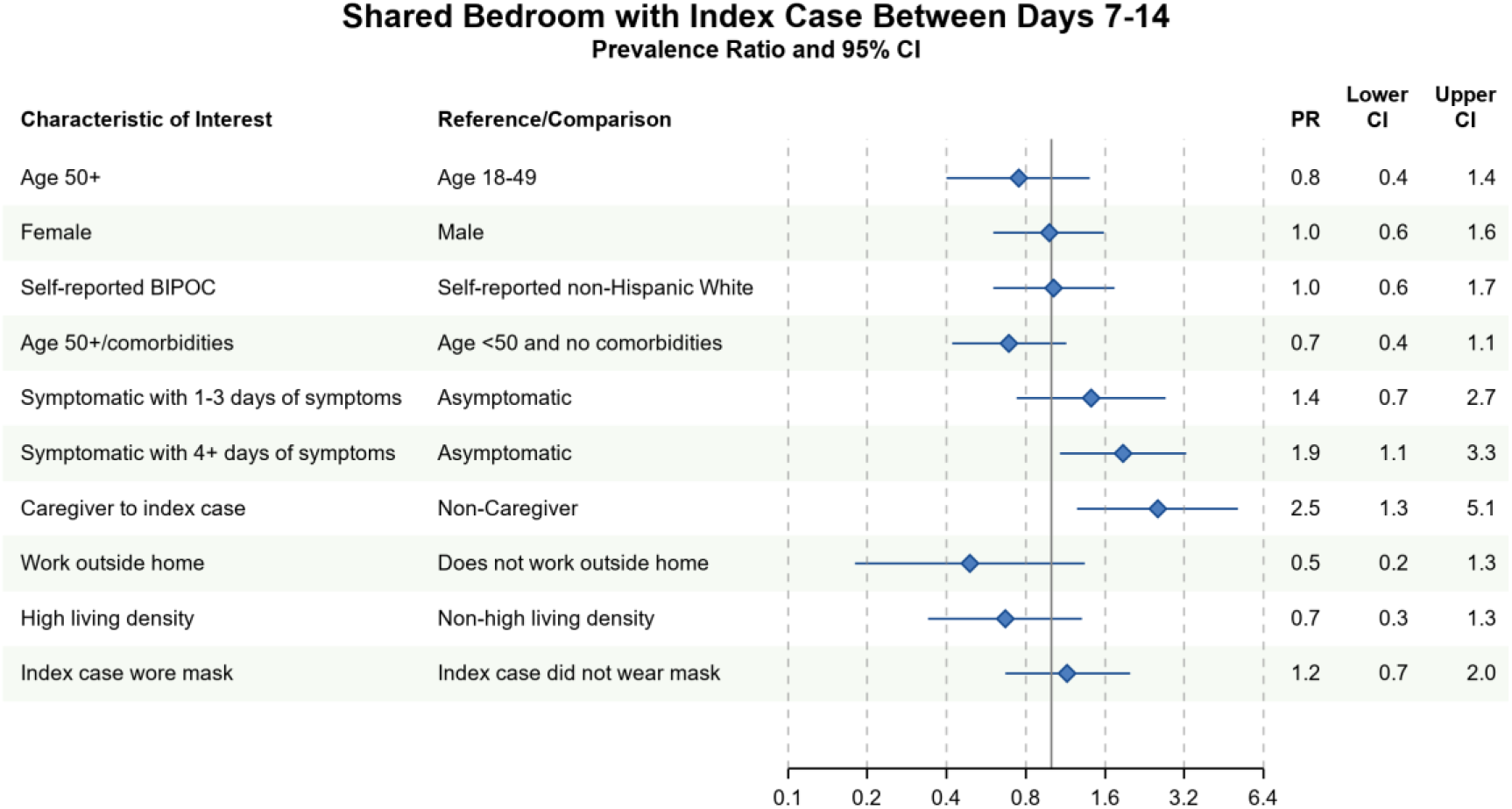
Bivariate sensitivity analyses of factors associated with sharing a bedroom with the index case at any time between Days 7-14 of cohort participation. Solid dots (PR) and solid lines (95% CI) display imputed estimates <u>using chained multiple imputation for clustered data</u>. PR and 95% CI graphed on transformed natural log scale. Vertical solid line denotes the null value of the PR (PR=1.0) on the exponentiated scale. X-axis labels correspond to the exponentiated scale. BIPOC=Black, Indigenous, People of Color. Table denotes the PR, lower 95% CI and upper 95% CI.

**Table S1.** Number and proportion of household contacts engaging in reported behaviors at study entry and Day 14. P-values were calculated using Yang’s test for changes between Day 0-14 on complete cases. 85 participants were missing ‘masking at home’ responses and 119 were evaluable. 41 participants were missing responses for all other variables and 163 participants were evaluable. See Figure 1 in main text.

| Behavior | N (%) Household Contacts Reporting at Cohort Entry | N (%) Household Contacts Reporting at Day 14 | P-value |
| --- | --- | --- | --- |
| Share kitchen | 148 (91) | 124 (76) | 0.003 |
| Eat together | 111 (68) | 90 (55) | 0.013 |
| Share bathroom | 105 (65) | 92 (56) | 0.066 |
| Watch TV together | 103 (63) | 89 (55) | 0.068 |
| Share car rides | 101 (62) | 66 (41) | <0.001 |
| Share electronic devices | 65 (40) | 54 (33) | 0.092 |
| Share bedroom | 58 (36) | 44 (27) | 0.015 |
| Masking at home | 28 (24) | 31 (26) | 0.614 |

**Table S2.**
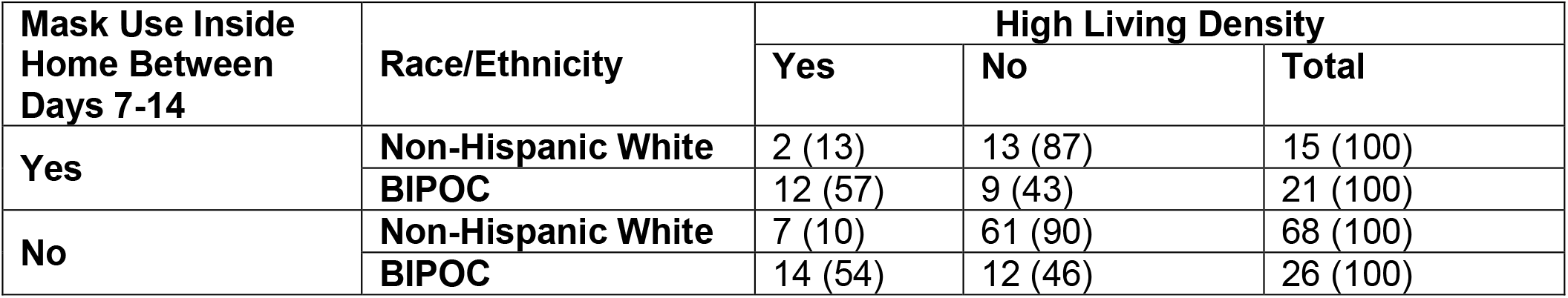
Number (row %) of household contacts living in high density households among self-reported race/ethnicity and masking behavior.

**Table S3.**
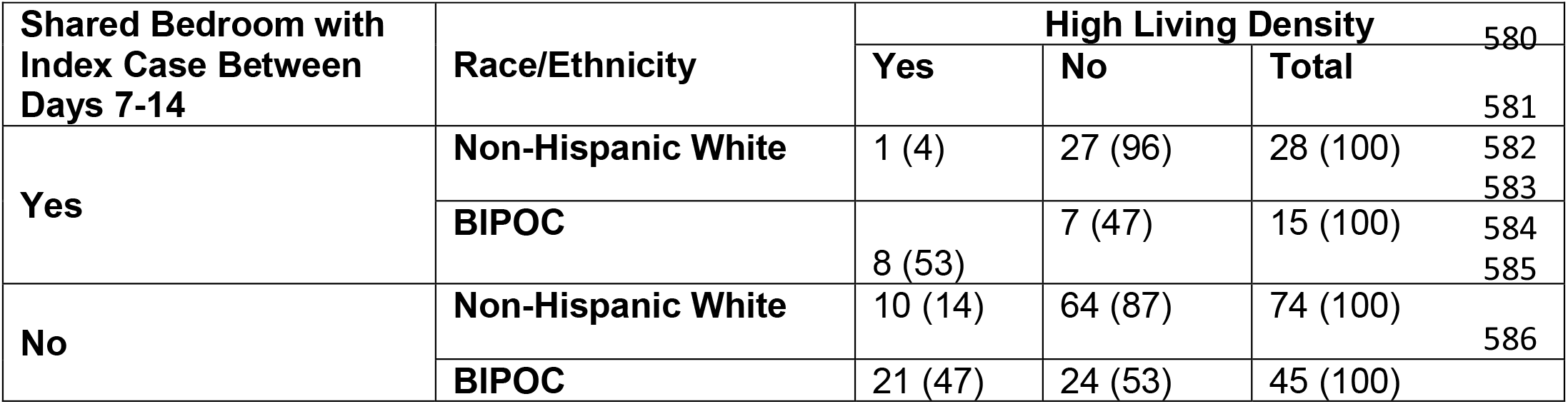
Number (%) of household contacts living in high density households among self-reported race/ethnicity and bedroom sharing with index case.

**Table S4.** Frequency and prevalence of household contacts who reported ever masking or ever sharing a bedroom with the index case between Days 7-14 of cohort participation.

| **Outcome:** | **Mask Use Inside the Home** |  | **Sharing a Bedroom** |  |
| --- | --- | --- | --- | --- |
| **Household-Contact Characteristics** | **N** | **Prevalence** | **N** | **Prevalence** |
| Age (restricted to 18 and older) |  |  |  |  |
| 18-49 (ref) | 16 | 0.26 | 24 | 0.33 |
| 50+ | 9 | 0.36 | 8 | 0.24 |
| Sex |  |  |  |  |
| Male (ref) | 15 | 0.26 | 20 | 0.27 |
| Female | 21 | 0.29 | 24 | 0.27 |
| Race/Ethnicity |  |  |  |  |
| Non-Hispanic White | 15 | 0.18 | 28 | 0.27 |
| Participants of Color | 21 | 0.45 | 16 | 0.26 |
| Aged 50 or older or any comorbidities<sup>a</sup> |  |  |  |  |
| No | 12 | 0.21 | 24 | 0.35 |
| Yes | 24 | 0.35 | 19 | 0.22 |
| Missing | 6 |  | 6 |  |
| Duration of COVID-19 Symptoms<sup>b</sup> |  |  |  |  |
| No symptoms | 20 | 0.18 | 18 | 0.16 |
| 1-3 days | 5 | 0.16 | 8 | 0.26 |
| 4 or more days | 7 | 0.22 | 12 | 0.38 |
| Missing | 28 |  | 28 |  |
| Caregiver to index case<sup>b</sup> (restricted to 18 and older) |  |  |  |  |
| No | 10 | 0.24 | 7 | 0.15 |
| Yes | 15 | 0.34 | 23 | 0.47 |
| Missing | 0 |  | 2 |  |
| Work outside home on most days<sup>b</sup>(restricted to 18 and older) |  |  |  |  |
| No | 22 | 0.32 | 29 | 0.33 |
| Yes | 3 | 0.17 | 3 | 0.16 |
| Live in household with high living density |  |  |  |  |
| No | 22 | 0.16 | 35 | 0.29 |
| Yes | 14 | 0.22 | 9 | 0.21 |
| Missing | 74 |  | 0 |  |
| Index case wore mask<sup>b</sup> (include all age groups) |  |  |  |  |
| No | 4 | 0.09 | 15 | 0.25 |
| Yes | 32 | 0.37 | 27 | 0.27 |
| Missing | 0 |  | 3 |  |

a We considered individuals aged 50 or older or those with at least one comorbidity to be at higher risk for severe COVID-19 infection. If the household contact was the only member of their household with higher risk for severe COVID-19 infection, they were placed in the reference group. Household contacts who resided in households in which some members were not enrolled in COHOST, and where there were no known household members over aged 50 or with comorbidities, were coded as missing.
b Between days 7-14 of cohort observation

## REFERENCES

1 He X, Lau EHY, Wu P, et al. Temporal dynamics in viral shedding and transmissibility of COVID-19. Nat Med 2020;26:672–5. doi:10.1038/s41591-020-0869-5

2 Tindale LC, Stockdale JE, Coombe M, et al. Evidence for transmission of COVID-19 prior to symptom onset. Elife 2020;9:e57149. doi:10.7554/eLife.57149

3 Benefield AE, Skrip LA, Clement A, et al. SARS-CoV-2 viral load peaks prior to symptom onset: a systematic review and individual-pooled analysis of coronavirus viral load from 66 studies. MedRxiv 20202028 [Preprint]. September 30, 2020 [cited 2021 Jun 01]. doi:10.1101/2020.09.28.20202028

4 Cerami C, Popkin-Hall ZR, Rapp T, et al. Household Transmission of Severe Acute Respiratory Syndrome Coronavirus 2 in the United States: Living Density, Viral Load, and Disproportionate Impact on Communities of Color. Clin Infect Dis 2021;ciab701. doi:10.1093/cid/ciab701

5 Grijalva CG, Rolfes MA, Zhu Y, et al. Transmission of SARS-COV-2 Infections in Households - Tennessee and Wisconsin, April-September 2020. MMWR Morb Mortal Wkly Rep 2020;69:1631–4. doi:10.15585/mmwr.mm6944e1

6 Sachdev DD, Chew Ng R, Sankaran M, et al. Contact tracing outcomes among household contacts of fully vaccinated COVID-19 patients - San Francisco, California, January 29-July 2, 2021. Clin Infect Dis 2021;ciab1042. doi:10.1093/cid/ciab1042

7 Jansen L, Tegomoh B, Lange K, et al. Investigation of a SARS-CoV-2 B.1.1.529 (Omicron) Variant Cluster — Nebraska, November–December 2021. MMWR Morb Mortal Wkly Rep 2021;70:1782–4. doi:10.15585/mmwr.mm705152e3

8 Baker JM, Nakayama JY, O’Hegarty M, et al. SARS-CoV-2 B.1.1.529 (Omicron) Variant Transmission Within Households - Four U.S. Jurisdictions, November 2021-February 2022. MMWR Morb Mortal Wkly Rep 2022;71:341–6. doi:10.15585/mmwr.mm7109e1

9 Julin CH, Robertson AH, Hungnes O, et al. Household Transmission of SARS-CoV-2: A Prospective Longitudinal Study Showing Higher Viral Load and Increased Transmissibility of the Alpha Variant Compared to Previous Strains. Microorganisms 2021;9:2371. doi:10.3390/microorganisms9112371

10 VoPham T, Weaver MD, Adamkiewicz G, et al. Social Distancing Associations with COVID-19 Infection and Mortality Are Modified by Crowding and Socioeconomic Status. Int J Environ Res Public Health 2021;18:4680. doi:10.3390/ijerph18094680

11 Martinez DA, Klein EY, Parent C, et al. Latino Household Transmission of Severe Acute Respiratory Syndrome Coronavirus 2. Clin Infect Dis 2021;ciab753. doi:10.1093/cid/ciab753

12 Achenbach J, Bever L, Booth W, et al. What you need to know about the coronavirus variants. Washington Post. 2021.https://www.washingtonpost.com/health/2021/11/29/covid-variants/ (Accessed 18 Jul 2022).

13 Sah P, Vilches TN, Moghadas SM, et al. Accelerated vaccine rollout is imperative to mitigate highly transmissible COVID-19 variants. EClinicalMedicine 2021;35:100865. doi:10.1016/j.eclinm.2021.100865

14 Mandavilli A. Reaching ‘Herd Immunity’ Is Unlikely in the U.S., Experts Now Believe. The New York Times. 2021.https://www.nytimes.com/2021/05/03/health/covid-herd-immunity-vaccine.html (Accessed 6 Jan 2022).

15 Kaiser Family Foundation. KFF COVID-19 Vaccine Monitor Dashboard | KFF. https://www.kff.org/coronavirus-covid-19/dashboard/kff-covid-19-vaccine-monitor-dashboard/ (Accessed 6 Jan 2022).

16 Yang Z, Sun X, Hardin JW. A note on the tests for clustered matched-pair binary data. Biom J 2010;52:638–52. doi:10.1002/bimj.201000035

17 Gopstein D. clust. bin. pair: Statistical Methods for Analyzing Clustered Matched Pair Data. 2016. https://CRAN.R-project.org/package=clust.bin.pair (Accessed 15 Feb 2021).

18 Quartagno M, Carpenter J. jomo: Multilevel Joint Modelling Multiple Imputation. 2022.https://CRAN.R-project.org/package=jomo (Accessed 31 May 2022).

19 Carpenter JR, Kenward MG. Multiple Imputation and its Application: Carpenter/Multiple Imputation and its Application. Chichester, UK: John Wiley & Sons, Ltd 2013.doi:10.1002/9781119942283

20 Denford S, Morton K, Horwood J, et al. Preventing within household transmission of Covid-19: is the provision of accommodation to support self-isolation feasible and acceptable? BMC Public Health 2021;21:1–13. doi:10.1186/s12889-021-11666-z

21 Oppel Jr RA, Gebeloff R, Lai KKR, et al. The Fullest Look Yet at the Racial Inequity of Coronavirus. The New York Times. 2020.https://www.nytimes.com/interactive/2020/07/05/us/coronavirus-latinos-african-americans-cdc-data.html (Accessed 18 Jul 2022).

22 Peck P. The Virus Is Showing Black People What They Knew All Along. The Atlantic. 2020.https://www.theatlantic.com/health/archive/2020/12/pandemic-black-death-toll-racism/617460/ (Accessed 18 Jul 2022).

23 Powdthavee N, Riyanto YE, Wong ECL, et al. When face masks signal social identity: Explaining the deep face-mask divide during the COVID-19 pandemic. PLoS One 2021;16:e0253195. doi:10.1371/journal.pone.0253195

24 Virginia Department of Health. Protect your health: masks. Coronavirus. https://www.vdh.virginia.gov/coronavirus/protect-yourself/masks/ (Accessed 27 Aug 2022).

25 Pei S, Yamana TK, Kandula S, et al. Burden and characteristics of COVID-19 in the United States during 2020. Nature 2021;598:338–41. doi:10.1038/s41586-021-03914-4

26 Najjar-Debbiny R, Gronich N, Weber G, et al. Effectiveness of Paxlovid in Reducing Severe Coronavirus Disease 2019 and Mortality in High-Risk Patients. Clin Infect Dis 2022;ciac443. doi:10.1093/cid/ciac443

27 Lamb YN. Nirmatrelvir Plus Ritonavir: First Approval. Drugs 2022;82:585–91. doi:10.1007/s40265-022-01692-5

28 Jørgensen SB, Nygård K, Kacelnik O, et al. Secondary Attack Rates for Omicron and Delta Variants of SARS-CoV-2 in Norwegian Households. JAMA 2022;327:1610. doi:10.1001/jama.2022.3780

29 Madewell ZJ, Yang Y, Longini IM, et al. Household Secondary Attack Rates of SARS-CoV-2 by Variant and Vaccination Status: An Updated Systematic Review and Meta-analysis. JAMA Netw Open 2022;5:e229317. doi:10.1001/jamanetworkopen.2022.9317

30 Antonelli M, Pujol JC, Spector TD, et al. Risk of long COVID associated with delta versus omicron variants of SARS-CoV-2. The Lancet 2022;399:2263–4. doi:10.1016/S0140-6736(22)00941-2

31 Yi C, Sun X, Ling Z, et al. Jigsaw puzzle of SARS-CoV-2 RBD evolution and immune escape. Cell Mol Immunol 2022;19:848–51. doi:10.1038/s41423-022-00884-z

